# Quality of life in hyperkalemia: baseline analysis of a cohort study of management of hyperkalemia in patients with chronic kidney disease or heart failure in Japan

**DOI:** 10.64898/2026.03.24.26349144

**Authors:** Ken-ei Sada, Hajime Yamazaki, Takafumi Wakita, Yosuke Yamamoto, Jui Wang, Yoshihiro Onishi, Tadateru Hamada, Ryotaro Ide, Masayoshi Takeda, Shunichi Fukuhara, Yugo Shibagaki

## Abstract

**Background:** Hyperkalemia is common in chronic kidney disease (CKD) and chronic heart failure (CHF), often leading to treatment dilemmas regarding renin-angiotensin-aldosterone system (RAAS) inhibitors. Although potassium binders and dietary restrictions are central to chronic management, their quality-of-life (QOL) impact remains insufficiently described. This study aimed to characterize real-world treatment patterns and evaluate treatment impact on QOL.

**Methods:** We analyzed baseline data from a prospective cohort in Japanese nephrology and cardiology outpatient clinics. Participants were adults with CKD (≥ stage G3) or CHF (New York Heart Association class II-IV) who initiated potassium binders within 6 months. Clinical data, serum potassium values, and patient-reported outcomes (generic QOL, disease/treatment-specific QOL, and adherence measures) were obtained at enrollment.

**Results:** Among 347 patients, the median age was 75 years, and 74% were male; 93% had CKD. At enrollment, 300 patients were receiving potassium binders, and 59% were prescribed a RAAS inhibitor. Dietary therapy was implemented in 29%. Physical scores of generic QOL were lower than population norms, whereas mental scores were comparable. Treatment-specific QOL scores indicated that potassium binders had a smaller impact on QOL than dietary therapy. Adherence to potassium binders was high.

**Conclusions:** Concurrent use of RAAS inhibitors and potassium binders was common, suggesting that binders may support RAAS inhibitor continuation. Potassium binders showed less perceived impact than dietary restrictions, indicating that pharmacologic potassium control may be acceptable to patients managing multiple lifestyle limitations. These findings highlight the role of potassium binders in maintaining both RAAS inhibitor therapy and QOL.

## Introduction

Hyperkalemia is a potentially life-threatening condition that not only causes muscle weakness and fatigue but may also lead to malignant arrhythmias such as ventricular fibrillation or even cardiac arrest [1]. Recently, hyperkalemia has attracted attention as a shared risk factor in patients with chronic kidney disease (CKD) and chronic heart failure (CHF) from the perspective of the cardiorenal connection. In these patients, impaired potassium excretion and metabolic abnormalities associated with disease progression frequently lead to hyperkalemia, making the appropriate management of hyperkalemia of great clinical importance [2]. In the acute phase of hyperkalemia, insulin–glucose therapy, calcium preparations, or diuretics are commonly used, whereas in the chronic phase, treatment mainly relies on dietary management (potassium intake restriction) and the administration of potassium binders.

Renin–angiotensin–aldosterone system (RAAS) inhibitors are widely used in the treatment of CKD and CHF for their renal and cardiac protective effects [3]. However, RAAS inhibitors are known to increase serum potassium levels and may cause hyperkalemia [4]. When RAAS inhibitor-induced hyperkalemia persists, discontinuation of these organ-protective therapies may be considered, potentially leading to worsening of long-term outcomes [5]. Hence, a dilemma: reduce or stop the RAAS inhibitor, or continue the RAAS inhibitor while putting patients on dietary restrictions/potassium binders?

Patients with chronic medical conditions often experience reduced health-related quality of life (HR-QOL) [6]. Likewise, patients who have both CHF or CKD and hyperkalemia may experience additional limitations on daily life arising from multiple sources, including physical and psychological distress from symptoms, complex medication regimens, dietary restrictions, and clinic visits. In such patients, not only disease severity but also treatment-related lifestyle restrictions are likely to contribute to diminished HR-QOL, as is the case in hyperkalemia management. However, studies focusing specifically on HR-QOL among patients with hyperkalemia remain limited.

Building on this background, we designed and conducted a prospective cohort study to elucidate real-world patterns of hyperkalemia management and to examine how treatment approaches and serum potassium control influence patients’ HR-QOL [7]. Overall HR-QOL was measured using a generic instrument, and HR-QOL specific to a disease or treatment was measured using a disease-specific instrument. The generic instrument to be used was the recently developed 8-Item QOL General (QGEN-8) [8]. The QGEN-8 is designed to provide scores relative to those in the general population. The disease-specific instrument to be used was the Quality of Life Disease Impact Scale-7 item (QDIS-7) [9]. The QDIS-7 not only measures the impact of a specified condition (e.g., a chronic disease and a treatment) on QOL, but also provides scores that can be compared between different conditions [10]. In this report, we address the first objective of the study – to describe treatment patterns and the impact of treatment on patients’ HR-QOL.

## Methods

### Study Design and Setting

This study was a 6-month, multicenter, prospective cohort study. Details have been published elsewhere [7]. The setting was the nephrology and cardiology subspecialty outpatient clinics in Japan. Data were collected from medical records and via patient-reported outcomes (PROs). In this report, we analyzed data collected at enrollment.

### Participants

Participants were aged 20 years or older, had either CKD (stage G3 or higher) or CHF (New York Heart Association class II–IV), were receiving potassium binder treatment, and had started a potassium binder no more than 6 months before enrollment or had a potassium binder scheduled at enrollment. Participants were consecutively enrolled between May 2022 and January 2024 at 35 nephrology and/or cardiology outpatient clinics in Japan.

### Data Analyzed

In this study, we analyzed the following data at enrollment: patient characteristics, serum potassium (sK) concentrations, use of hyperkalemia-related treatments and dietary therapy, and data obtained via PROs, including generic QOL, disease-or treatment-specific QOL, and medication adherence to potassium binders.

#### Patient-reported outcomes

We analyzed a total of 8 PROs, (1) to (8) below. To assess generic QOL, we used the QGEN-8 [8]. It is designed to provide norm-based scores, which are scores relative to those of a representative sample of the general population. These general population scores are set to have a mean of 50 and a standard deviation (SD) of 10, with higher scores indicating a favorable QOL. We analyzed the two summary scores obtained from the QGEN-8, asking about the past month: (1) the physical component and (2) the mental component. Although the source study also collected MOS Short-Form 12-Item Health Survey (SF-12) and 36-item survey (SF-36) data, we reported only the QGEN-8 scores, as we confirmed that the scores from these instruments provided essentially the same information as the QGEN-8. QOL specific to underlying disease (CKD or CHF), potassium binder treatment, and dietary therapy were assessed using the QDIS-7. The QDIS-7 is an attribute-specific rather than disease-specific measure, allowing comparisons of the impact of different attributes – such as diseases, exposures, symptoms, and treatments – across individuals or within the same individual [10]. The validity of the QDIS-7 in patients with hyperkalemia has been demonstrated in a preceding study [11]. The QDIS-7 consists of 7 items that assess the impact of a specific attribute on QOL over the past 1 month [9]. For example, one item asks, “In the past month, how many times did [CONDITION] limit your physical activities such as walking or climbing stairs?” In this study, three versions were used, substituting [CONDITION] with (3) “chronic kidney disease” or “heart failure”, (4) “treatment with potassium binder”, or (5) “dietary therapy”, where (3) was administered to all participants, while (4) and (5) were administered only to patients receiving their respective treatment. Similar to QGEN-8, QDIS-7 scores are standardized to have a mean of 50 and an SD of 10 [9]. However, the target population for standardization is not the general population but individuals with one or more predefined chronic medical conditions [10]. Unlike QGEN-8, higher QDIS-7 scores indicate greater impact of the disease or treatment on QOL, that is, an unfavorable state. Adherence to potassium binder treatment was assessed by three instruments [7]: (6) the compliance with prescriptions scale for potassium binder (CWP) was a 14-item scale assessing behaviors associated with correct potassium binder medication use over the past one month; (7) the medicine-taking behavior scale (MTB) was a 3-item scale assessing reluctance to take medication over the past one month; (8) the overall adherence single-item measure (OAD) was a single question asking the degree of taking the prescribed potassium binder over the past one month. For (7) and (8), the sum of the item scores (1 to 5 points for each) was converted to a score ranging from 0 to 100 (best to worst). The score range for (8) is 1-5 (worst to best).

### Statistical Analysis

This study focused on describing cross-sectional data at enrollment. We described patient characteristics, the status of serum potassium, and the implementation of major treatments that affect serum potassium. We also summarized the breakdown of potassium binder prescriptions and the proportion of dietary therapy administration. We described the distributions of scores of QGEN-8 (physical and mental components), QDIS-7 (for CKD/CHF, for potassium binder, and for dietary therapy), and adherence to potassium binder prescriptions (CWP, MTB, and OAD).

To explore characteristics of patients reporting any impact (in at least one item out of seven) on QDIS-7 for potassium binder, we performed stratified analyses by several attributes of patient characteristics, treatments, and PRO scores other than QDIS-7 scores for potassium binder. The other PRO scores were categorized by the median or a nearby, easily quantifiable score.

All analyses were conducted on a complete-case basis. Data were summarized as means and SDs, medians and interquartile ranges (IQRs), or frequencies and proportions. Where appropriate, histograms were used to visualize data distributions. As this study did not have specific hypotheses, no statistical tests were performed. Instead, standardized mean differences [12] were used when assessing the magnitude of differences in distributions of a specific variable across strata or in distributions of two variables. The magnitude of the effect size (SMD: magnitude of difference) was interpreted as: small, SMD⍰= ⍰0.2; medium, SMD⍰= ⍰0.5; or large, SMD⍰= ⍰0.8 [12]. SAS 9.4 (Cary, NC, USA) was used for analyses.

## Results

### Patients

A total of 352 patients were enrolled. After excluding 2 patients who did not meet eligibility criteria and 3 who withdrew consent, 347 patients were included in the analysis. The characteristics of these patients are shown in Table 1. The median (IQR) age was 75 (66 to 81) years, and 74% were male. The underlying disease was CKD and HF in 93% and 7% of the patients, respectively.

**Table 1.** Characteristics of patients included in the analysis.

|  | Number of subjects |  |
| --- | --- | --- |
|  | N=347 |  |
| Age [years] | 75 | [66, 81] |
| Sex (male) | 74% | (256) |
| Body mass index<sup>1)</sup> [kg/m<sup>2</sup>] | 24.1 | [21.7, 26.5] |
| Underlying disease |  |  |
| Chronic kidney disease | 93% | (322) |
| Stage 3b | 24% | (85) |
| Stage 4 | 42% | (147) |
| Stage 5 | 26% | (90) |
| Duration of disease [years] | 4.0 | [1.5, 8.0] |
| Heart failure (HF) | 7% | (25) |
| NYHA-II | 6% | (21) |
| NYHA-III | 1% | (3) |
| NYHA-IV | 0% | (1) |
| Duration of disease [years] | 1.0 | [0.5, 7.7] |
| Comorbidities/history |  |  |
| Diabetes mellitus | 52% | (182) |
| Hypertension | 95% | (328) |
| Cardiovascular diseases |  |  |
| Atrial fibrillation | 12% | (40) |
| Stroke | 9% | (31) |
| Arrhythmia (atrial or ventricular) | 7% | (23) |
| Myocardial infarction | 5% | (19) |
| Unstable angina pectoris | 5% | (16) |
| Peripheral arterial disease | 4% | (15) |
| Valvular heart disease | 4% | (14) |

Values are median [interquartile range] or proportion (frequency).
1) Missing in 21 patients.
NYHA, New York Heart Association functional classification.

Of the 347 patients, 300 had already been prescribed a potassium binder at enrollment, while 47 started a potassium binder immediately after enrollment. The mean (SD) serum potassium (sK) at enrollment in each subgroup was 4.81 (0.56) mmol/L and 5.35 (0.48) mmol/L, respectively.

### Treatments associated with hyperkalemia

Table 2 summarizes the proportion of patients receiving major treatments associated with hyperkalemia. RAAS inhibitors were prescribed to 59% of the patients. Beta-blockers and non-steroidal anti-inflammatory drugs (NSAIDs) were used in 24% and 13% of the patients, respectively. Treatments that may lower sK included loop diuretics and sodium bicarbonate, prescribed to 30% and 16% of the patients, respectively. Of the 300 patients who had already been prescribed potassium binders at enrollment, RAAS inhibitors were used in 168 (58%). At enrollment, 58% (n = 202) received calcium polystyrene sulfonate, 16% (n = 54) received sodium polystyrene sulfonate, and 26% (n = 91) received sodium zirconium cyclosilicate, where data at the first potassium binder prescription were used for the patients enrolled before potassium binder initiation. Regarding dietary interventions, potassium restriction was implemented in 14% of patients (n = 47). Some form of dietary restriction, including energy restriction, salt restriction, protein restriction, phosphorus restriction, or potassium restriction, was implemented in 29% of patients (n = 99).

**Table 2.** Prescriptions of drugs potentially affecting serum potassium at enrollment.

|  |  | Number of subjects |
| --- | --- | --- |
|  |  | N=347 |
| **Drugs that may increase serum potassium** |  |  |
| Renin-angiotensin-aldosterone system inhibitors <sup>1)</sup> | 59% | (204) |
| Angiotensin-converting enzyme inhibitors | 3% | (11) |
| Angiotensin II receptor blockers | 47% | (163) |
| Angiotensin receptor neprilysin inhibitors | 3% | (12) |
| Mineralocorticoid receptor antagonists | 5% | (18) |
| β-blockers | 24% | (84) |
| Non-steroidal anti-inflammatory drugs | 13% | (46) |
| **Drugs that may lower serum potassium** |  |  |
| Loop diuretics (oral) | 30% | (103) |
| Sodium bicarbonate (oral) | 16% | (56) |
| Thiazide diuretics | 7% | (23) |
| Glucose with insulin | 2% | (7) |
| Calcium gluconate | 1% | (3) |
| Loop diuretics (intravenous) | 1% | (2) |
| Sodium bicarbonate (intravenous) | 1% | (2) |

1) When data were limited to only the 300 patients who were already prescribed potassium binders at enrollment, 168 of them (58%) were prescribed renin-angiotensin-aldosterone system inhibitors.

### Patient-reported outcomes

#### Generic quality of life

Generic QOL scores using the QGEN-8 were obtained for 343 of the 347 patients. Table 3 summarizes generic QOL scores grouped by patient feature. In the entire cohort, the physical component score was lower than the national average of 50 points, while the mental component score was comparable to the national average. By patient characteristics, the physical component score was lower among older patients and women. There was no notable difference in QOL scores between the 47 patients without a potassium binder prescription (pre-initiation) and the 296 patients already receiving a potassium binder. Likewise, no difference in QOL scores was observed between the 98 patients who received dietary therapy and the 245 who did not.

**Table 3.** Quality of Life at enrollment, measured by the 8-Item QOL General (QGEN-8)

|  |  | Physical component | Mental component |
| --- | --- | --- | --- |
| Overall | (n = 343) | 43.7 [6.7] | 50.5 [6.0] |
| Age |  |  |  |
| <75 | (n = 163) | 46.2 [6.2] | 50.6 [5.8] |
| ≥75 | (n = 180) | 41.5 [6.4] | 50.5 [6.1] |
| Sex |  |  |  |
| Male | (n = 253) | 44.5 [6.9] | 50.6 [5.8] |
| Female | (n = 90) | 41.4 [5.7] | 50.3 [6.4] |
| Underlying disease |  |  |  |
| Chronic kidney disease | (n = 318) | 44.0 [6.6] | 50.6 [5.9] |
| Stage 3b | (n = 85) | 44.7 [6.9] | 50.9 [6.5] |
| Stage 4 | (n = 144) | 44.3 [6.6] | 51.0 [5.7] |
| Stage 5 | (n = 89) | 42.8 [6.3] | 49.6 [5.7] |
| Heart failure | (n = 25) | 40.1 [7.1] | 49.8 [6.4] |
| NYHA-II | (n = 21) | 41.1 [7.2] | 50.3 [6.1] |
| NYHA-III | (n = 3) | 34.7 [3.5] | 43.3 [4.5] |
| NYHA-IV | (n = 1) | 34.8 [.] | 58.1 [.] |
| Potassium binder prescription <sup>1)</sup> |  |  |  |
| No | (n = 47) | 43.3 [7.1] | 49.2 [6.3] |
| Yes | (n = 296) | 43.8 [6.7] | 50.7 [5.9] |
| Dietary therapies |  |  |  |
| No | (n = 245) | 43.6 [6.8] | 50.9 [6.0] |
| Yes | (n = 98) | 44.0 [6.6] | 49.6 [5.9] |

Values are means [standard deviations]. The scores were missing in four patients.
1) Potassium binders were first prescribed before and after enrollment in 300 and 47 patients, respectively.
NYHA, New York Heart Association functional classification.

#### Disease (treatment) impact on quality of life

Figure 1 shows the 3 versions of QDIS-7 scores. Among the 347 patients, CKD/CHF impact scores were obtained in 342 individuals. Of these, 100 individuals (29%) had scores greater than the standard value of 50 points. The mean score (SD) was 47.2 (6.0). Among the 300 patients receiving potassium binder at enrollment, QDIS-7 scores for potassium binder were obtained in 296 individuals. Among these, 23 (8%) had scores greater than the standard value of 50 points. The mean score (SD) was 43.3 (4.0). The lowest possible score (no impact reported for any of the 7 questions) was observed in 170 patients (57%), while 126 patients (43%) reported some impact. Among the 99 patients receiving dietary therapy at enrollment, QDIS-7 data on dietary therapy were obtained for 90. Of these, 28 patients (31%) had scores greater than the standard value of 50 points. The mean score (SD) was 47.5 (5.8).

**Figure 1.**
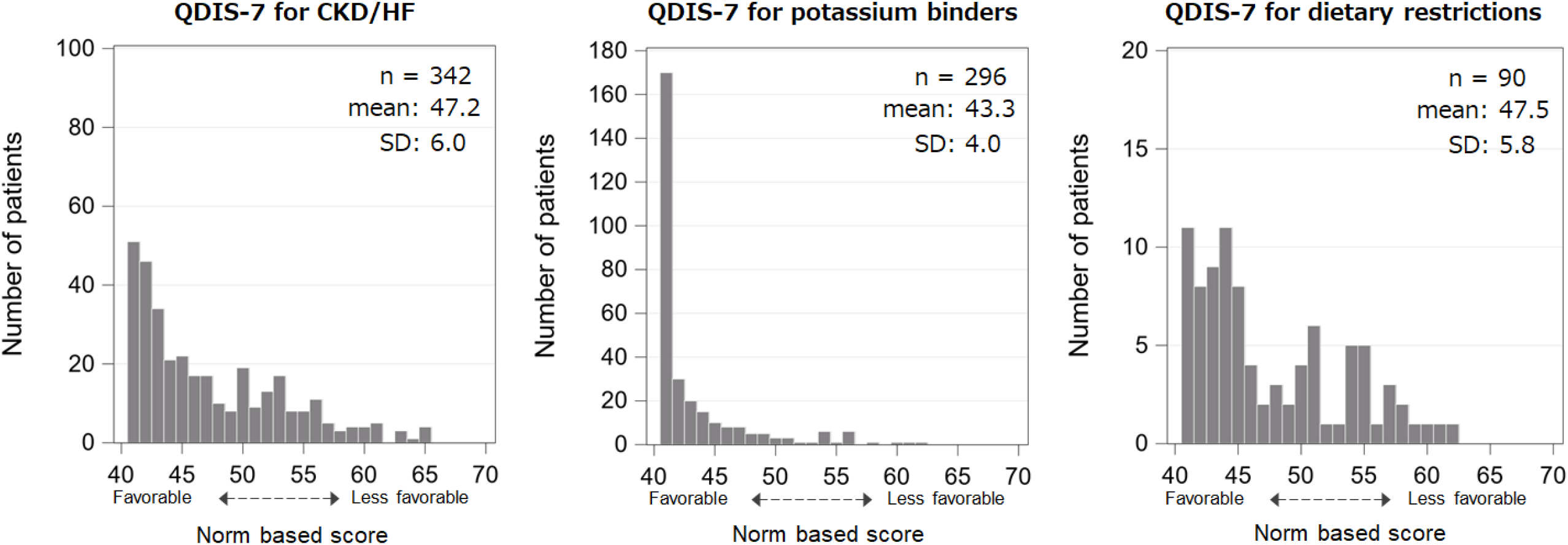
Distribution of the Quality-of-Life Disease Impact Scale-7 item (QDIS-7) scores indicating the impact of disease and treatment on QOL at registration. QDIS-7 scores are standardized based on the distribution of scores among the general population with chronic diseases (mean 50, standard deviation 10). Higher values indicate a greater impact (worse condition). The lowest possible score is 41.1, which indicates no problems reported for any of the questions comprising QDIS-7

The difference between the QDIS-7 for potassium binder and that for dietary therapy was an SMD of 0.84, corresponding to a “large difference.”

#### Adherence to potassium binders

Figure 2 shows the distribution of PRO scores assessing adherence to potassium binder prescriptions. In CWP (n = 296), 53% of patients reported the lowest possible score (no problems reported in any of 14 questions). In MTB (n = 297), 49% of patients reported the lowest possible score. In OAD (n = 294), 82% of patients reported the best response, “5. Took all doses.”

**Figure 2.**
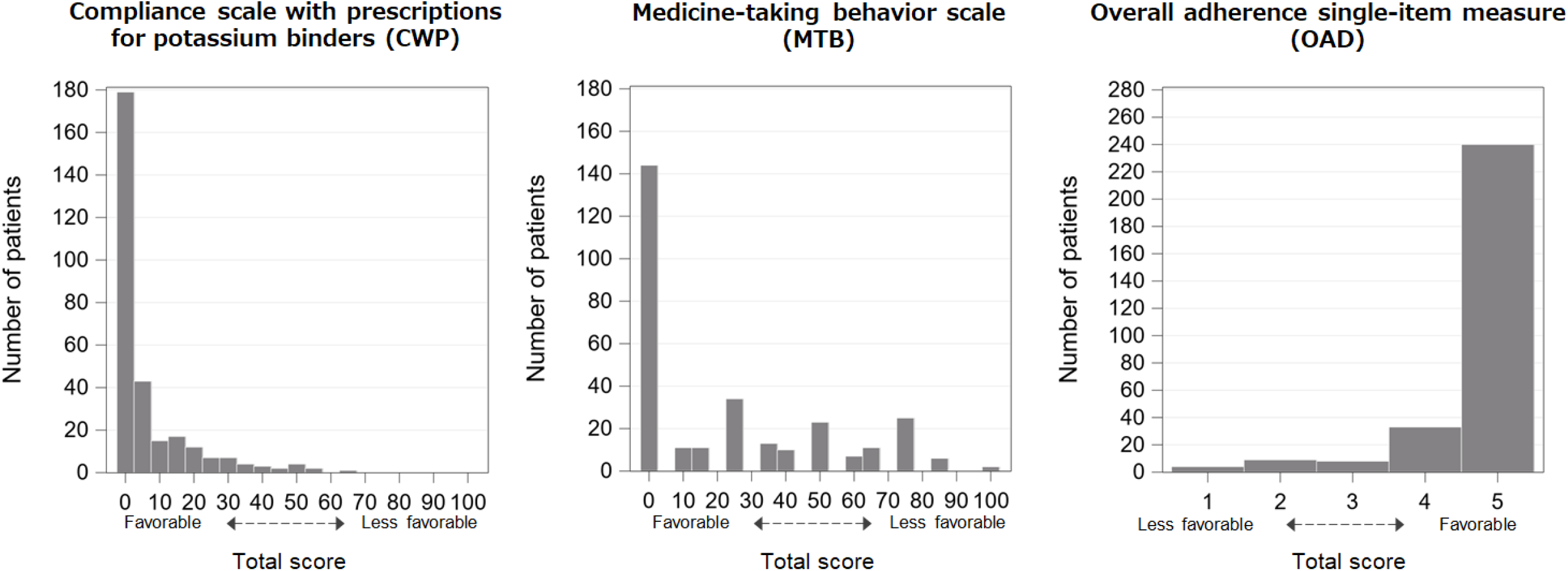
Distribution of patient-reported outcome (PRO) scores for adherence to potassium binders at enrollment. The scores presented are simple total scores. In the two types of scores on the left, a score of 0 indicates no problems reported for any of the questions comprising that PRO

#### Factors associated with the impact of potassium binder treatments

Figure 3 shows the associations of patient characteristics or PRO scores with the QDIS-7 for potassium binders. The proportion of patients reporting some impact was greater in the groups aged 74 years or younger, female, with more advanced CKD, CHF, and polypharmacy. Correlations with generic PROs were more pronounced, especially for the mental component scores.

**Figure 3.**
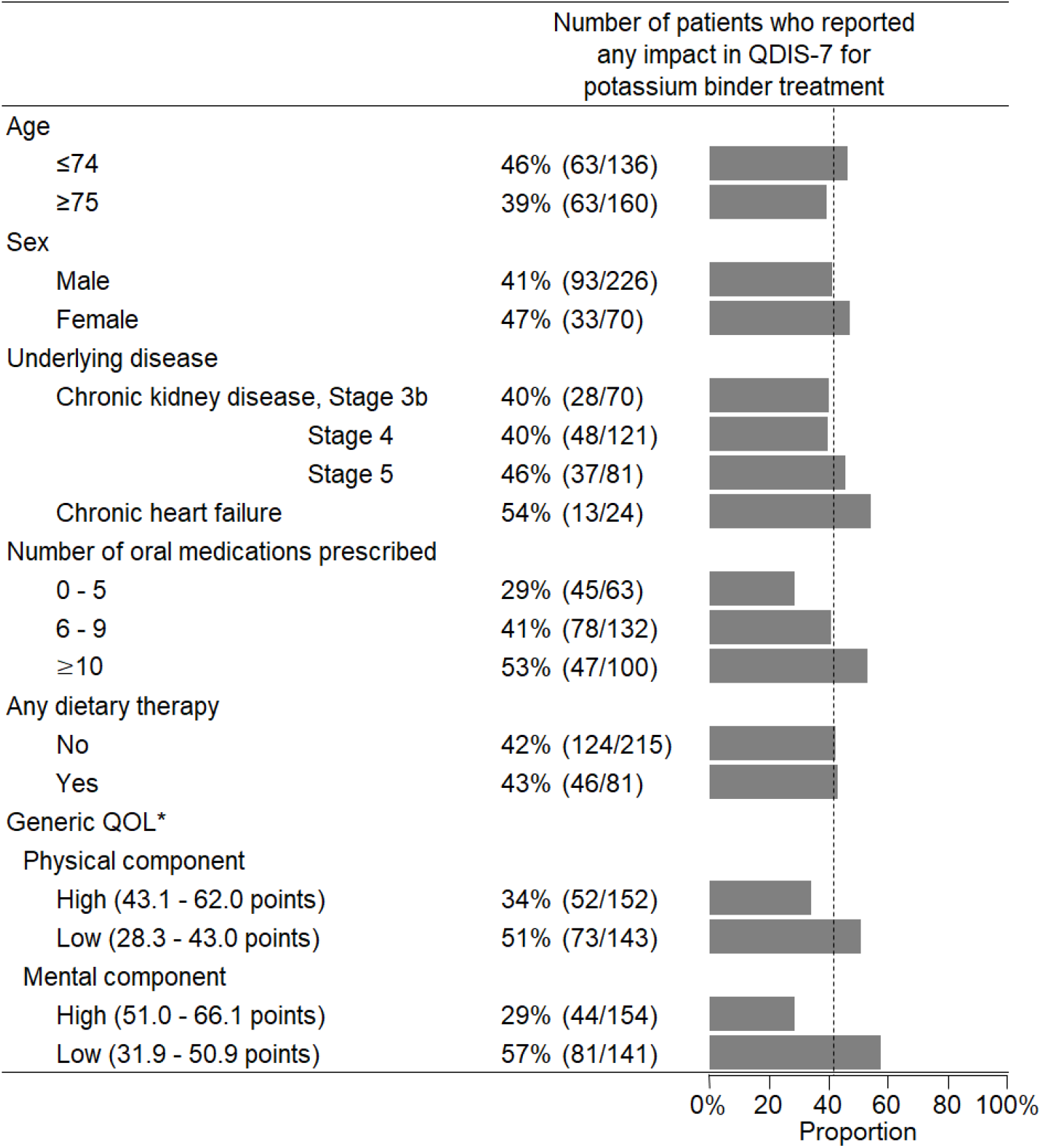
Exploration of factors potentially associated with the reported impact of potassium binder treatment via the Quality-of-Life Disease Impact Scale-7 item (QDIS-7). The dotted line indicates the proportion of 42.6% in all patients. * Measured by the 8-Item QOL General (QGEN-8)

## Discussion

In this study, we described treatment patterns and the impact of treatment on patients’ HR-QOL. Among enrolled patients, 93% had CKD, and RAAS inhibitors were prescribed in as much as 59% in total and 58% in potassium binder users at enrollment. The treatment impact on QOL assessed by QDIS-7 was considerably smaller for potassium binders compared with dietary restriction. More than half (57%) of potassium binder users did not report any constraints from a potassium binder treatment on QDIS-7 questions. Adherence to potassium binders was favorable overall.

In our cohort, approximately 60% of patients receiving potassium binders were concurrently treated with a RAAS inhibitor. It is generally preferable to continue RAAS inhibitors in patients with CKD or CHF because of their organ-protective effects, but their use is often limited by hyperkalemia. In this study, the higher proportion of RAAS inhibitor use compared with a prior hyperkalemia cohort [5] suggests that potassium binders may have facilitated the continuation of RAAS inhibitor therapy. Previous reports have shown that discontinuation of a RAAS inhibitor is associated with lower use of potassium binders [5,13], indicating that potassium binder therapy may help maintain RAAS inhibitor treatment in clinical practice. Given that discontinuation of RAAS inhibitors due to hyperkalemia has been suggested to worsen patient outcomes, and in line with the cross-specialty expert consensus on the optimal management of hyperkalemia [14], strategies such as dietary potassium restriction or the use of potassium binders to enable continued RAAS inhibition may be advantageous.

If active treatment of hyperkalemia is considered important for enabling the continuation of RAAS inhibitor therapy, the next issue to consider is the treatment burden it imposes on patients. This study also suggested that potassium binders may have less constraint on patients’ QOL than dietary therapy. Among patients with CKD, approximately 35% have been reported to experience moderate to high treatment burden, which was negatively correlated with QOL [15]. Similarly, in patients with CHF, about 20% experienced a high level of treatment burden, which was also inversely associated with QOL [16]. However, these studies did not clarify which specific aspects of treatment contributed most to the perceived burden. In CHF, dietary sodium restriction is an essential component of management, yet achieving recommended targets in intervention studies has often proven difficult [17]. For CKD, dietary therapy is even more complex, involving not only sodium restriction but also limitations on protein and phosphorus intake [18]. Recent Kidney Disease Improving Global Outcomes (KDIGO) guidelines [19] further emphasize the need for individualized dietary adaptations regarding sodium, phosphorus, potassium, and protein intake tailored to patients’ needs, CKD severity, and comorbid conditions, often requiring specialized input from renal dietitians or accredited nutrition providers. Under such restrictive conditions, additional potassium restriction may impose a considerable impact on both CKD and CHD patients.

The QDIS-7 is a disease-specific QOL measure that enables comparison of disease impacts across different conditions and treatment factors, which generic QOL scales or conventional disease-specific QOL scales cannot reveal [10]. In fact, the QDIS-7 was reported to have higher validity than the generic QOL scale (SF-36) for measuring the burden on QOL by CKD according to its stages (severity) [11]. In our analysis, no difference in the generic QOL scale (QGEN-8) was observed between those with and without dietary therapy; however, the use of QDIS-7 enabled us to detect differences in treatment-related impacts. Taken together, these findings suggest that patients may prefer potassium control through oral potassium binders rather than further intensification of dietary restrictions. The use of potassium binders may not only facilitate the continuation of RAAS inhibitor therapy but also help maintain patients’ QOL by allowing some relaxation of dietary restrictions. Several limitations should be acknowledged. First, the study’s cross-sectional design does not allow assessment of temporal patterns in the concomitant use of potassium binders and RAAS inhibitors. It remains unclear how these therapies have been adjusted over time. Second, the QOL assessment was conducted at baseline, and thus, the impact of potassium binder therapy may not yet have been fully captured. In contrast, the assessment may already reflect the impact of other dietary restrictions beyond potassium limitation. These issues warrant further evaluation in the ongoing longitudinal analysis.

In conclusion, this study characterized real-world treatment patterns and HR-QOL in patients with hyperkalemia. In this cohort, the proportion of RAAS inhibitor use among patients receiving potassium binders was relatively high, although the cross-sectional design does not allow assessment of how these therapies were adjusted over time. QOL scores suggested that potassium binders were associated with a smaller perceived impact than dietary therapy, indicating that pharmacologic potassium control may be more acceptable to patients already facing multiple dietary restrictions. These findings imply that potassium binders may help support continued RAAS inhibitor therapy (and broaden patients’ food and drink options) while minimizing additional treatment impacts on QOL.

Longitudinal studies are required to clarify treatment trajectories and QOL changes over time.

## Data Availability

No data are available. However, data requests can be sent to the authors.

## Acknowledgements

We wish to express our appreciation to AstraZeneca K.K. and the Patient Driven Academic League (PeDAL) for their project management efforts, and to A2 Healthcare Corporation for their data collection efforts. We also appreciate the cooperation of the patients and facilities participating in this study. Christopher Holmes of the Institute for Health Outcomes & Process Evaluation Research provided comments, suggestions, and editorial advice on an earlier version of the manuscript.

Instrument availability: QDIS-7 is available without royalty for academic scholarly researchers. For Japan-only projects, users register through. For all other projects, users register through <https://eprovide.mapi-trust.org/>.

## Funding

This study was funded by AstraZeneca K.K.

## Compliance with Ethical Standards

### Conflicts of interest

**Employment:** Tadateru Hamada, Ryotaro Ide, and Masayoshi Takeda (AstraZeneca K.K.),

**Consultancies:** Yosuke Yamamoto (Nippon Shinyaku Co.),

**Honoraria:** Ken-ei Sada (AstraZeneca K.K.), Yugo Shibagaki (AstraZeneca K.K., Otsuka Pharma Co., and Kyowa Kirin Co.), Hajime Yamazaki (Janssen Pharmaceutical K.K., Mitsubishi Tanabe Pharma, Kowa Company Limited, AstraZeneca K.K., and Takeda Pharmaceutical Company Limited.), Yosuke Yamamoto (Sun Pharma, Asahi Kasei Pharma, Toray Industries, Inc., and Ono Pharmaceutical Co., Ltd.), Unrelated to the submitted work, consultation fees for Hajime Yamazaki and Shunichi Fukuhara were paid to Kyoto University under contracts with Takeda Pharmaceutical Co., Ltd. and Magmitt Pharmaceutical Co., Ltd.

### Ethical approval

All procedures performed in studies involving human participants were in accordance with the ethical standards of the institutional and/or national research committee at which the studies were conducted (as listed below) and with the 1964 Helsinki declaration and its later amendments or comparable ethical standards.

### List of ethics committees and approval numbers

Azumino Red Cross Hospital Ethics Committee, R04-A-02; Ethics Committee of Kasugai Municipal Hospital, 22-7; Komatsu Ishikawa Central Hospital Ethics Committee, R4-6; Non-Profit Organization MINS Research Ethics Committee, MINS-REC-220204; Hiroshima City Hiroshima Citizens Hospital Ethics Review Committee, 2022-25; Saga Memorial Hospital Ethics Committee, 9/3/2022; Ethics Committee on Epidemiological and its related Studies, Sakuragaoka Campus, Kagoshima University, 220154; Clinical Ethics Committee of New Tokyo Hospital, 0274-1 and 0274-3; Nagaoka Chuo General Hospital Ethics Committee, 541; The Research Ethics Review Committee of NHO Nagoya Medical Center, 2022-024; The Ethics Review Committee of Toyama City General Hospital, 2022-07.

### Informed consent

Informed consent was obtained from all individual participants included in the study.

### Study registration

The study was registered with ClinicalTrials.gov (NCT05297409).

## Notes

### Author Declarations

Ethics committees listed below gave ethical approval for this work: Azumino Red Cross Hospital Ethics Committee, Ethics Committee of Kasugai Municipal Hospital, Komatsu Ishikawa Central Hospital Ethics Committee, Non-Profit Organization MINS Research Ethics Committee, Hiroshima City Hiroshima Citizens Hospital Ethics Review Committee, Saga Memorial Hospital Ethics Committee, Ethics Committee on Epidemiological and its related Studies, Sakuragaoka Campus, Kagoshima University, Clinical Ethics Committee of New Tokyo Hospital, Nagaoka Chuo General Hospital Ethics Committee, The Research Ethics Review Committee of NHO Nagoya Medical Center, The Ethics Review Committee of Toyama City General Hospital.

### Summary of Updates

Corrected a spelling error in one author's name.

## References

1. Hunter RW, Bailey MA. Hyperkalemia: pathophysiology, risk factors and consequences. Nephrol Dial Transplant. 2019;34:iii2–11.

2. Wang AY-M. Optimally managing hyperkalemia in patients with cardiorenal syndrome. Nephrol Dial Transplant. 2019;34:iii36–44.

3. Tsukamoto S, Uehara T, Azushima K, Wakui H, Tamura K. Updates for cardio-kidney protective effects by angiotensin receptor-neprilysin inhibitor: Requirement for additional evidence of kidney protection. J Am Heart Assoc. 2023;12:e029565.

4. Kovesdy CP, Appel LJ, Grams ME, Gutekunst L, McCullough PA, Palmer BF, et al. Potassium homeostasis in health and disease: A scientific workshop cosponsored by the National Kidney Foundation and the American Society of Hypertension. J Am Soc Hypertens. 2017;11:783–800.

5. Kashihara N, Kohsaka S, Kanda E, Okami S, Yajima T. Hyperkalemia in real-world patients under continuous medical care in Japan. Kidney Int Rep. 2019;4:1248–60.

6. Fukuhara S, Yamazaki S, Hayashino Y, Green J. Measuring health-related quality of life in patients with end-stage renal disease: why and how. Nat Clin Pract Nephrol. 2007;3:352–3.

7. Shibagaki Y, Yamazaki H, Wakita T, Ware JE, Wang J, Onishi Y, et al. Impact of treatment of hyperkalaemia on quality of life: design of a prospective observational cohort study of long-term management of hyperkalaemia in patients with chronic kidney disease or chronic heart failure in Japan. BMJ Open. 2023;13:e074090.

8. Ware JE Jr. Improved Items for Estimating SF-36 Profile and Summary Component Scores: Construction and Validation of an 8-Item QOL General (QGEN) Survey. Med Care. 2025;63:300–310.

9. Ware JE Jr, Gandek B, Guyer R, Deng N. Standardizing disease-specific quality of life measures across multiple chronic conditions: development and initial evaluation of the QOL Disease Impact Scale (QDIS®). Health Qual Life Outcomes. 2016;14:84.

10. Fukuhara S, Green J, Wakita T, Yamamoto Y, Yamazaki H, Ware JE Jr. The QDIS-7: using one scale to measure the disease-specific quality-of-life impact of different medical conditions. Sci Rep. 2025;15:21756.

11. Fukuhara S, Yamazaki H, Wakita T, Ware JE Jr, Wang J, Onishi Y, et al. Validation of a new instrument for measuring disease-specific quality of life: A pilot study among patients with chronic kidney disease and hyperkalemia. Ann Clin Epidemiol. 2023;5:13–9.

12. Cohen J. Statistical power analysis for the behavioral sciences. 2. Hillsdale: L. Erlbaum Associates; 1988.

13. Pollack CV Jr, Arroyo D, Kanda E, Lázaro IJS, Lesén E, Franzén S, et al. Duration of sodium zirconium cyclosilicate treatment and continuation of RAASi therapy after a hyperkalaemia episode. ESC Heart Fail. 2025;12:1776–85.

14. Kitai T, Maruyama S, Kuwahara K, Tamura K, Kinugawa K, and Kashihara N. Establishing cross-specialty expert consensus on the optimal management of hyperkalemia in patients with heart failure and chronic kidney disease. Circ J. 2025;89:470–478.

15. Al-Mansouri A, Al-Ali FS, Hamad AI, Mohamed Ibrahim MI, Kheir N, Ibrahim RA, et al. Assessment of treatment burden and its impact on quality of life in dialysis-dependent and pre-dialysis chronic kidney disease patients. Res Social Adm Pharm. 2021;17:1937–44.

16. Nordfonn OK, Morken IM, Bru LE, Larsen AI, Husebø AML. Burden of treatment in patients with chronic heart failure - A cross-sectional study. Heart Lung. 2021;50:369–74.

17. Burgermaster M, Rudel R, Seres D. Dietary sodium restriction for heart failure: A systematic review of intervention outcomes and behavioral determinants. Am J Med. 2020;133:1391–402.

18. Palmer SC, Maggo JK, Campbell KL, Craig JC, Johnson DW, Sutanto B, et al. Dietary interventions for adults with chronic kidney disease. Cochrane Database Syst Rev. 2017;4:CD011998.

19. Kidney Disease: Improving Global Outcomes (KDIGO) CKD Work Group. KDIGO 2024 Clinical Practice Guideline for the Evaluation and Management of Chronic Kidney Disease. Kidney Int. 2024;105:S117–S314.

